# Viral cultures, PCR Cycle threshold values and viral load estimation for COVID-19 infectious potential assessment in transplant patients: systematic review - Protocol Version 30 December 2021

**DOI:** 10.1101/2021.12.30.21268509

**Authors:** Tom Jefferson, Elizabeth A. Spencer, Susanna Maltoni, Jon Brassey, Igho J. Onakpoya, Elena C. Rosca, Annette Plüddemann, David H. Evans, John M. Conly, Carl J. Heneghan

## Abstract

This is the protocol for a systematic review focussing on people receiving solid organ or hematopoietic stem cell transplants.

Our research questions are as follows:

What is the relationship between serial PCR Ct value or other measures of viral burden, and the likelihood and duration of the presence of infectious virus from viral culture, among transplant recipients with SARS-CoV-2 infection?

What is the influence of age, sex, underlying pathologies, degree of immunosuppression, vaccination status, COVID-19 symptoms and COVID-19 disease course on viral burden and the likelihood of presence of infectious SARS-CoV-2?

We will include single studies reporting serial Cts from sequential rt-PCR testing or other measures of viral burden such as RNA gene copies of respiratory samples (from nasopharyngeal specimens) along with viral culture data on the same samples, from patients about to receive a transplant or who are post transplant with SARS-CoV-2 infection.

## Background

Effective prevention of transmission of SARS-CoV-2 infections includes identifying potentially infectious individuals. Previous work has demonstrated the importance of careful description of cases and their follow up based on the close correlation between symptom onset, surrogates of viral load such as real-time PCR cycle threshold (rt-PCR Ct) and likelihood of shedding replication competent virus^1-4^. However this likelihood may vary across populations and settings^5^. The infectious dose in humans is unknown but in susceptible animal models such as the Syrian hamster it has been shown to be as low as five infectious particles^6^.

In people receiving immunosuppressive agents (henceforth “immunosuppressed”) either preceding or following a solid organ or hematopoietic stem cell transplant, infection with SARS-CoV-2 is likely to be of longer duration than in non-immunocompromised patients ^2, 7^. As infectious virus carriage may be greatly prolonged in immunosuppressed transplant patients compared to immune competent people, safe removal from isolation should be based on the potential for forward transmission of infectious virus as established using viral culture methods or suitable surrogate markers. Where this is not possible, care protocols may rely on substitute indicators such as rt-PCR Ct values or other estimates of viral load.

This review is part of a series aiming: a) to collate and evaluate the published data on potential infectivity assessment in immunosuppressed patients infected with SARS-CoV-2, including only studies employing viral cell culture, and b) to estimate the relationship between viral culture and repeat rt-PCR Ct values or other measures of viral burden among immunosuppressed patients. This review will focus on people receiving solid organ or hematopoietic stem cell transplants. Future reviews will focus on other immunosuppressed populations either because of underlying primary or secondary immunodeficiencies (e.g. HIV, primary hypogammaglobulinaemia, use of chemotherapy and other immunosuppressive drugs.

The series of reviews will focus on narrowing uncertainty around the prognostic significance for infectivity of surrogate viral burden measures.

## Methods

### Search Strategy

The following electronic databases will be searched: LitCovid, medRxiv, Google Scholar and the WHO Covid-19 database.

The literature search terms will be:

(coronavirus OR covid-19 OR SARS-CoV-2) AND (immunosuppressed OR immunocompromised OR transplant OR immunosuppression OR “immune deficient” OR HIV) AND (CPE OR “cytopathic effect” OR “Viral culture” OR “virus culture” OR vero OR “virus replication” OR “viral replication” OR “cell culture” or “viral load” OR “viral threshold” OR “log copies” OR “cycle threshold”). Each database will be searched from November 2019 until December 31, 2021. No language restrictions will be imposed.

### Screening

Screening will be performed independently by four researchers (JB, TJ, SM and ES). Title and abstract screening will identify studies for consideration of full text. Full text screening will be performed in duplicate and disagreements arbitrated by a third reviewer.

### Inclusion criteria

Studies reporting serial Cts from sequential rt-PCR testing or other measures of viral burden such as RNA gene copies of respiratory samples (from nasopharyngeal – NP - specimens) along with viral culture data on the same samples, from patients about to receive a transplant or who are post transplant with SARS-CoV-2 infection, will be included. Any primary study type will be considered for inclusion, providing it reports sufficient relevant information and detail. Studies reporting only in poster or abstract form will be excluded. Reviews will be excluded but the reference lists screened for potential relevant primary studies.

### Exclusion criteria

Studies using post-mortem samples only and non-respiratory samples only will be excluded. We will also exclude studies that did not perform viral cultures.

### Data extraction

Data will be extracted by one researcher and independently checked by a second; disagreements will be arbitrated by a third reviewer. Data will be extracted on study characteristics including population, setting, sampling and laboratory methods, clinical information, prescribed drugs, vaccination status, laboratory findings and clinical outcomes.

### Quality assessment

Quality of included studies will be assessed according to five criteria:

1. Were the criteria for diagnosing a case clearly reported and appropriate?
2. Was the reporting of patient/population characteristics including clinical symptoms, treatments with degree of immunosuppression and outcomes adequate?
3. Was the study period, including follow-up, sufficient to adequately assess any potential relationship between viral burden measures and likelihood of producing replication-competent virus and the rise in neutralizing antibodies?
4. Were the methods used to obtain rt-PCR results replicable, generalisable and appropriate? We consider that each study should establish the relationship between their Ct values and the target gene copy number, using internal standards^8^. Studies using internal controls can allow some degree of cross-calibration between the different PCR assays used.
5. Were the methods used to obtain viral culture results replicable and appropriate? We consider the methods used should, at a minimum, include a description of specimen sampling and management, preparation, media and cell line used, exclusion of contamination or co-infection (use of good controls and appropriate antibiotics and antimycotics and possible use of gene sequencing if available) and results of inspection of culture.

It is anticipated that for this relatively uncommon patient group, most studies reporting relevant data will be case studies or case series. These types of studies may be inherently biased in the selection of study participants, and might not fully represent the transplant patient population overall. However, they probably represent the most detailed longitudinal reports of the findings available.

### Data reporting and pooling

We will report study flow according to PRISMA reporting standards for systematic reviews and meta-analyses^9^. We will report study characteristics including age, sex, clinical symptoms, treatments and events in the participants in tabular form. We will present data on disease burden measures and on viral culture in tabular form. For studies reporting more than one patient participant, data will be extracted related to all transplant patient participants. We are not planning to pool the data to provide a cut off because the differences in laboratory practices and assays and sometimes the absence of internal controls while undertaking PCR are likely to impede meaningful comparisons^8, 10^. We plan instead to review narratively with a summary estimate of the relationship between viral burden and likelihood of producing replication-competent SARS-CoV-2 from NP specimens^2^.

If data are available and sufficient, the influence of age and/or sex on the relationship will be investigated. The results of these analyses may be presented in tables or figures, or solely in the text.

## Data Availability

All data produced in the present work will be contained in the manuscript

## Conflict of interest statements

TJ’s competing interests are accessible at: https://restoringtrials.org/competing-interests-tom-jefferson

CJH holds grant funding from the NIHR, the NIHR School of Primary Care Research, the NIHR BRC Oxford and the World Health Organization for a series of Living rapid review on the modes of transmission of SARs-CoV-2 reference WHO registration No2020/1077093. He has received financial remuneration from an asbestos case and given legal advice on mesh and hormone pregnancy tests cases. He has received expenses and fees for his media work including occasional payments from BBC Radio 4 Inside Health and The Spectator. He receives expenses for teaching EBM and is also paid for his GP work in NHS out of hours (contract Oxford Health NHS Foundation Trust). He has also received income from the publication of a series of toolkit books and for appraising treatment recommendations in non-NHS settings. He is Director of CEBM and is an NIHR Senior Investigator.

DE holds grant funding from the Canadian Institutes for Health Research and Li Ka Shing Institute of Virology relating to the development of Covid-19 vaccines as well as the Canadian Natural Science and Engineering Research Council concerning Covid-19 aerosol transmission. He is a recipient of World Health Organization and Province of Alberta funding which supports the provision of BSL3-based SARS-CoV-2 culture services to regional investigators. He also holds public and private sector contract funding relating to the development of poxvirus-based Covid-19 vaccines, SARS-CoV-2-inactivation technologies, and serum neutralization testing.

JMC holds grants from the Canadian Institutes for Health Research on acute and primary care preparedness for COVID-19 in Alberta, Canada and was the primary local Investigator for a *Staphylococcus aureus* vaccine study funded by Pfizer for which all funding was provided only to the University of Calgary. He is co-investigator on a WHO funded study using integrated human factors and ethnography approaches to identify and scale innovative IPC guidance implementation supports in primary care with a focus on low-resource settings and using drone aerial systems to deliver medical supplies and PPE to remote First Nations communities during the COVID-19 pandemic. He also received support from the Centers for Disease Control and Prevention (CDC) to attend an Infection Control Think Tank Meeting. He is a member and Chair of the WHO Infection Prevention and Control Research and Development Expert Group for COVID-19 and a member of the WHO Health Emergencies Programme (WHE) Ad-hoc COVID-19 IPC Guidance Development Group, both of which provide multidisciplinary advice to the WHO and for which no funding is received and from which no funding recommendations are made for any WHO contracts or grants. He is also a member of the Cochrane Acute Respiratory Infections Working Group.

JB is a major shareholder in the Trip Database search engine (www.tripdatabase.com) as well as being an employee. In relation to this work Trip has worked with a large number of organisations over the years, none have any links with this work. The main current projects are with AXA and SARS-CoV-2 (WHO Registration 2020/1077093-0) and is part of the review group carrying out rapid reviews for Collateral Global. He worked on Living rapid literature review on the modes of transmission of SARS-CoV-2 and a scoping review of systematic reviews and meta-analyses of interventions designed to improve vaccination uptake (WHO Registration 2021/1138353-0).

ECR was a member of the European Federation of Neurological Societies (EFNS) / European Academy of Neurology (EAN) Scientist Panel, Subcommittee of Infectious Diseases (2013 to 2017). Since 2021, she is a member of the International Parkinson and Movement Disorder Society (MDS) Multiple System Atrophy Study Group, the Mild Cognitive Impairment in Parkinson Disease Study Group, and the Infection Related Movement Disorders Study Group. She was an External Expert and sometimes Rapporteur for COST proposals (2013, 2016, 2017, 2018, 2019) for Neurology projects. She is a Scientific Officer for the Romanian National Council for Scientific Research.

AP holds grants from the NIHR School of Primary Care Research.

IJO and EAS have no interests to disclose.

SM is a pharmacist working for the Italian National Health System since 2002 and a member of one of the three Institutional Review Boards of Emilia-Romagna Region (Comitato Etico Area Vasta Emilia Centro) since 2018.

## References

1. Jefferson, T., et al., Viral cultures for COVID-19 infectious potential assessment – a systematic review. Clinical Infectious Diseases, 2020. 10.1093/cid/ciaa1764.

2. Rhee, C., et al., Duration of SARS-CoV-2 Infectivity: When is it Safe to Discontinue Isolation? Clinical Infectious Diseases, 2020. 10.1093/cid/ciaa1249.

3. Cevik, M., et al., SARS-CoV-2, SARS-CoV-1 and MERS-CoV viral load dynamics, duration of viral shedding and infectiousness: a living systematic review and meta-analysis. medRxiv, 2020: p. 2020.07.25.20162107. 10.1101/2020.07.25.20162107.

4. Tom, M.R. and M.J. Mina, To Interpret the SARS-CoV-2 Test, Consider the Cycle Threshold Value. Clinical Infectious Diseases, 2020. 10.1093/cid/ciaa619.

5. Jefferson, T., et al., Transmission of Severe Acute Respiratory Syndrome Coronavirus-2 (SARS-CoV-2) from pre and asymptomatic infected individuals. A systematic review. Clinical Microbiology and Infection, 2021. https://doi.org/10.1016/j.cmi.2021.10.015.

6. Rosenke, K., et al., Defining the Syrian hamster as a highly susceptible preclinical model for SARS-CoV-2 infection. Emerg Microbes Infect, 2020. 9(1): p. 2673–2684. 10.1080/22221751.2020.1858177.

7. Rajakumar, I.A.-O., et al., Extensive environmental contamination and prolonged severe acute respiratory coronavirus-2 (SARS CoV-2) viability in immunosuppressed recent heart transplant recipients with clinical and virologic benefit with remdesivir. (1559-6834 (Electronic)).

8. Evans, D., et al., The Dangers of Using Cq to Quantify Nucleic Acid in Biological Samples: A Lesson From COVID-19. Clinical Chemistry, 2021. 10.1093/clinchem/hvab219.

9. Page, M.J., et al., The PRISMA 2020 statement: an updated guideline for reporting systematic reviews. BMJ, 2021. 372: p. n71. 10.1136/bmj.n71.

10. IDSA and AMP joint statement on the use of SARS-CoV-2 PCR cycle threshold (Ct) values for clinical decision-making. 2021; Available from: https://www.idsociety.org/globalassets/idsa/public-health/covid-19/idsa-amp-statement.pdf.

